# Climate change and tuberculosis: an analytical framework

**DOI:** 10.1101/2025.02.18.25322451

**Authors:** Matthew J Saunders, Delia Boccia, Palwasha Y Khan, Lara Goscè, Antonio Gasparrini, Rebecca A Clark, Julia M Pescarini, Richard G White, Rein MGJ Houben, Matteo Zignol, Nebiat Gebreselassie, C Finn McQuaid

## Abstract

Climate change is likely to exacerbate a range of determinants which drive tuberculosis, the world’s leading infectious disease killer. However, tuberculosis is often neglected in wider climate health discussions.

Commissioned by the World Health Organization, we developed an analytical framework outlining potential causal relationships between climate change and tuberculosis. We drew on existing knowledge of tuberculosis determinants, identified which are likely to be sensitive to the effects of climate change, and conceptualised the mechanistic pathways through which this might occur. We collated evidence for these pathways through literature reviews. Our reviews found no studies directly linking climate change and tuberculosis, warranting research to build evidence for action.

The available evidence supports the existence of plausible links between climate change and tuberculosis, and highlights the need to include tuberculosis in climate risk adaptation and mitigation programmes, and climate-resilient funding and response mechanisms. Further evidence is urgently needed to quantify the effects of climate change on tuberculosis.

## Introduction

The health effects of climate change operate through complex and interconnected pathways, as outlined in the World Health Organization (WHO) Framework on Climate Change and Health, and further characterised in the Sixth Assessment Report of the Intergovernmental Panel on Climate Change (IPCC) and *Lancet* Countdown on climate change and health(1–3). Briefly, the WHO Framework postulates that climate-related hazards (e.g. extreme weather events, sea level rise) interact with vulnerabilities (e.g. gender, comorbidities) and exposures (e.g. food and health systems) leading to direct and indirect health effects. These include injuries and mortality; increases in zoonoses, food-, water-and vector-borne diseases, non-communicable diseases; and mental ill health(4). Climate change is also already affecting determinants of health by driving poverty, causing migration and displacement, worsening food and water insecurity, and disrupting access to healthcare and support systems(1–3). Importantly, many of the effects of climate change are cascading and compounding, and disproportionately affect populations in low- and middle-income countries where resilience and ability to adapt are lower(1–3).

These climate-sensitive determinants of health significantly overlap with key determinants of the global tuberculosis (TB) epidemic(5). Despite progress, TB continues to rank among the world’s leading causes of death(6). In 2023 alone, an estimated 10.8 million people fell ill from TB, and 1.25 million lost their lives to the disease(6). Determinants which heighten exposure to *Mycobacterium tuberculosis*, the causative agent of TB, such as overcrowding and poor living conditions, increase the risk of transmission and subsequent infection (hereafter referred to as TB infection). Meanwhile, determinants more specifically linked to health, which principally impair the immune system, such as undernutrition, HIV, alcohol use disorders, smoking, and diabetes, increase the risk of progressing to symptomatic and/or infectious disease (referred to here as TB disease, to distinguish it from TB infection)(6) and may worsen health outcomes. Many of these determinants can further exacerbate the known financial and psychosocial burden associated with TB, pushing TB-affected households deeper into poverty.

Notably, many countries with a high TB burden, including India, Indonesia and the Philippines(6), are also highly vulnerable to the effects of climate change, as measured by several recognised indices(7–9)(Figure 1). Positioning TB in the context of climate change has, however, been overlooked due to insufficient research. A recent scoping review suggested that climate change increases TB infection and disease risk, particularly among vulnerable populations(10), while a second review described the potential effects of climate change on several of the TB determinants described above(11). Despite these findings, the available evidence has not been systematically mapped against a comprehensive framework describing the potential pathways linking climate change and TB. Although some frameworks have been proposed (12,13), further work is needed to establish a global, consensus-based framework on the basis of evidence. Consequently, the current and potential future effects of climate change on TB are often overlooked in the wider climate and health discussion(14,15), and no coherent strategy exists to support countries to mitigate these effects. Developing a framework is therefore an urgent priority to guide effective policy and action.

**Figure 1.**
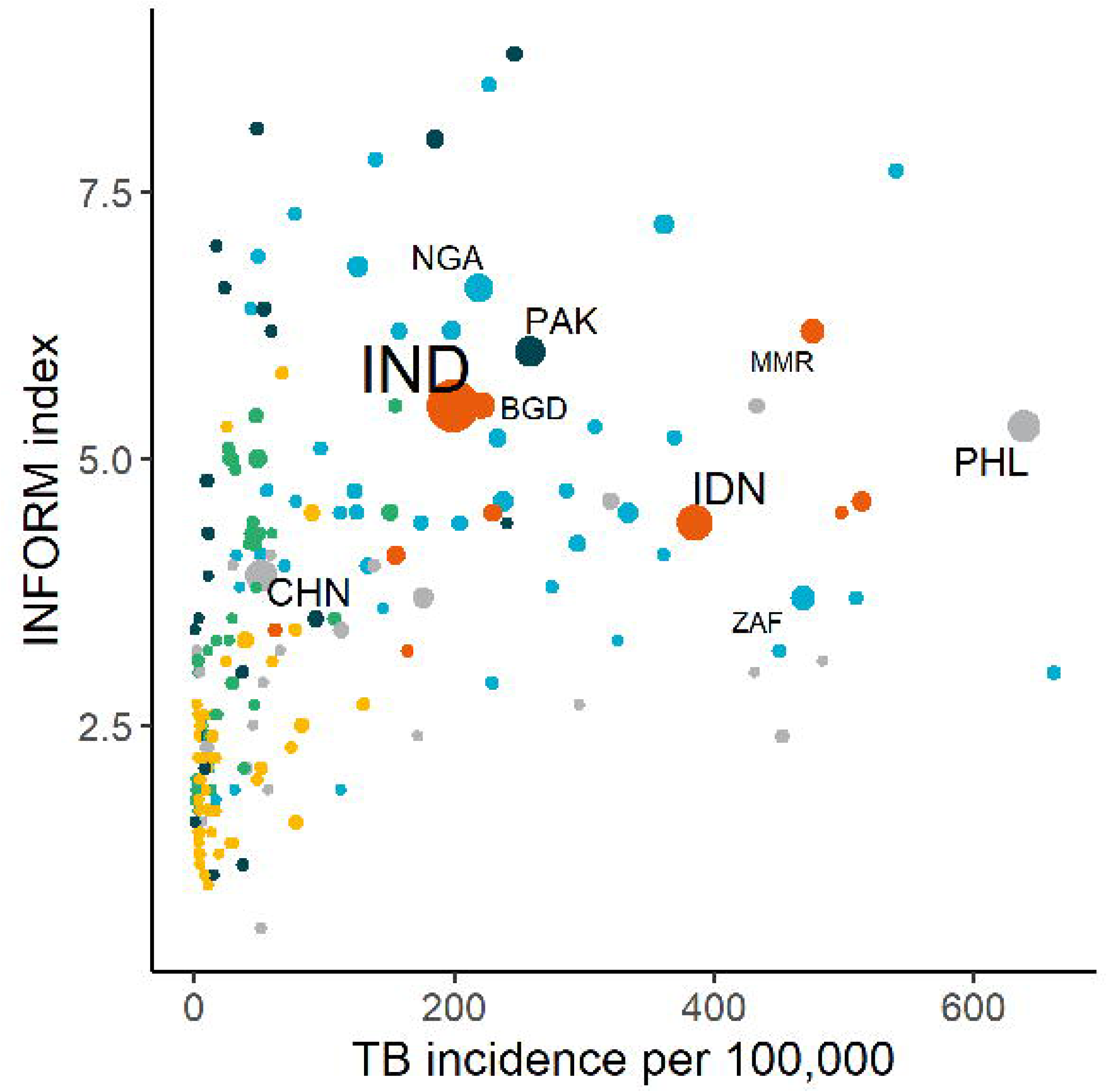
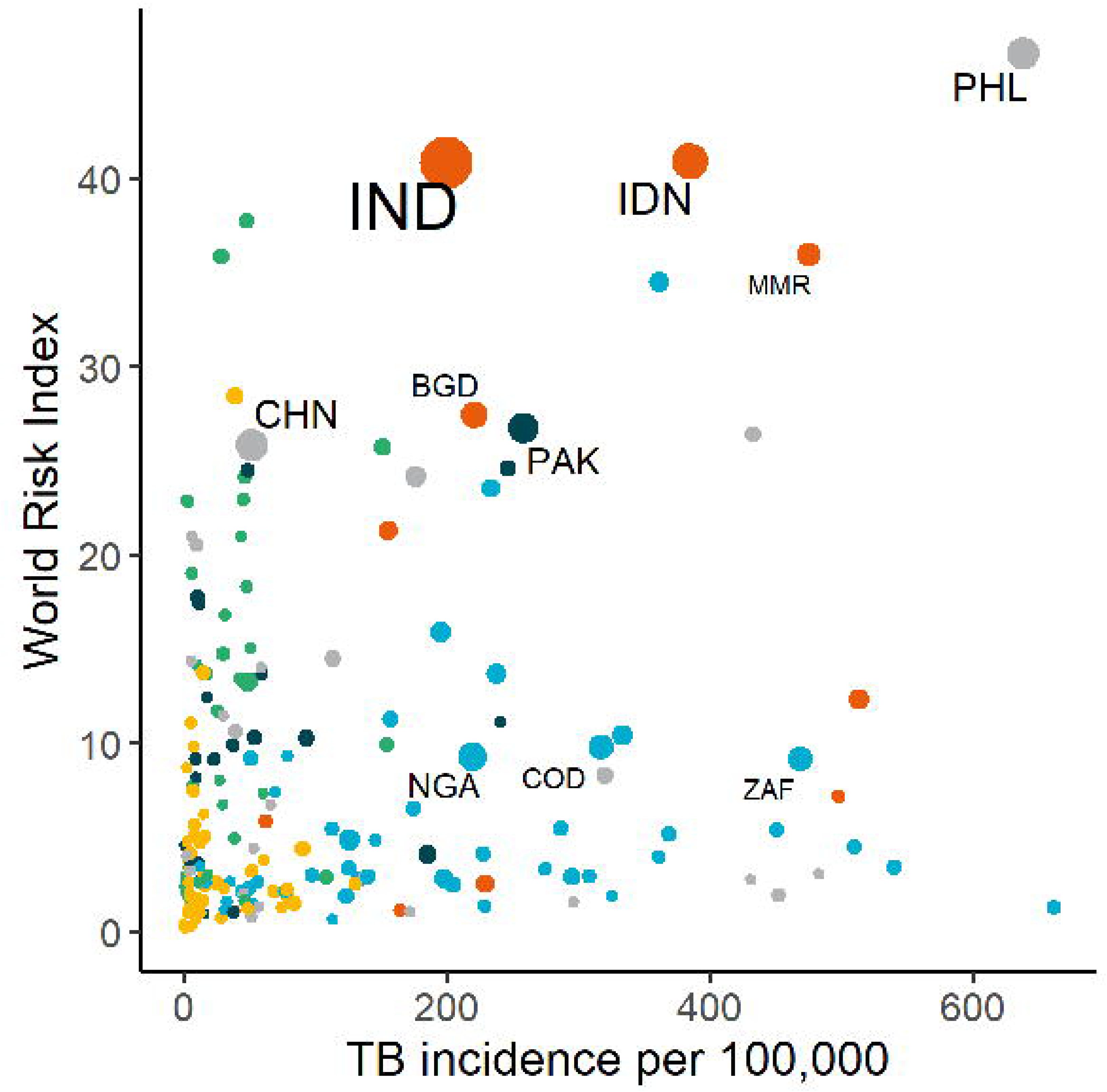
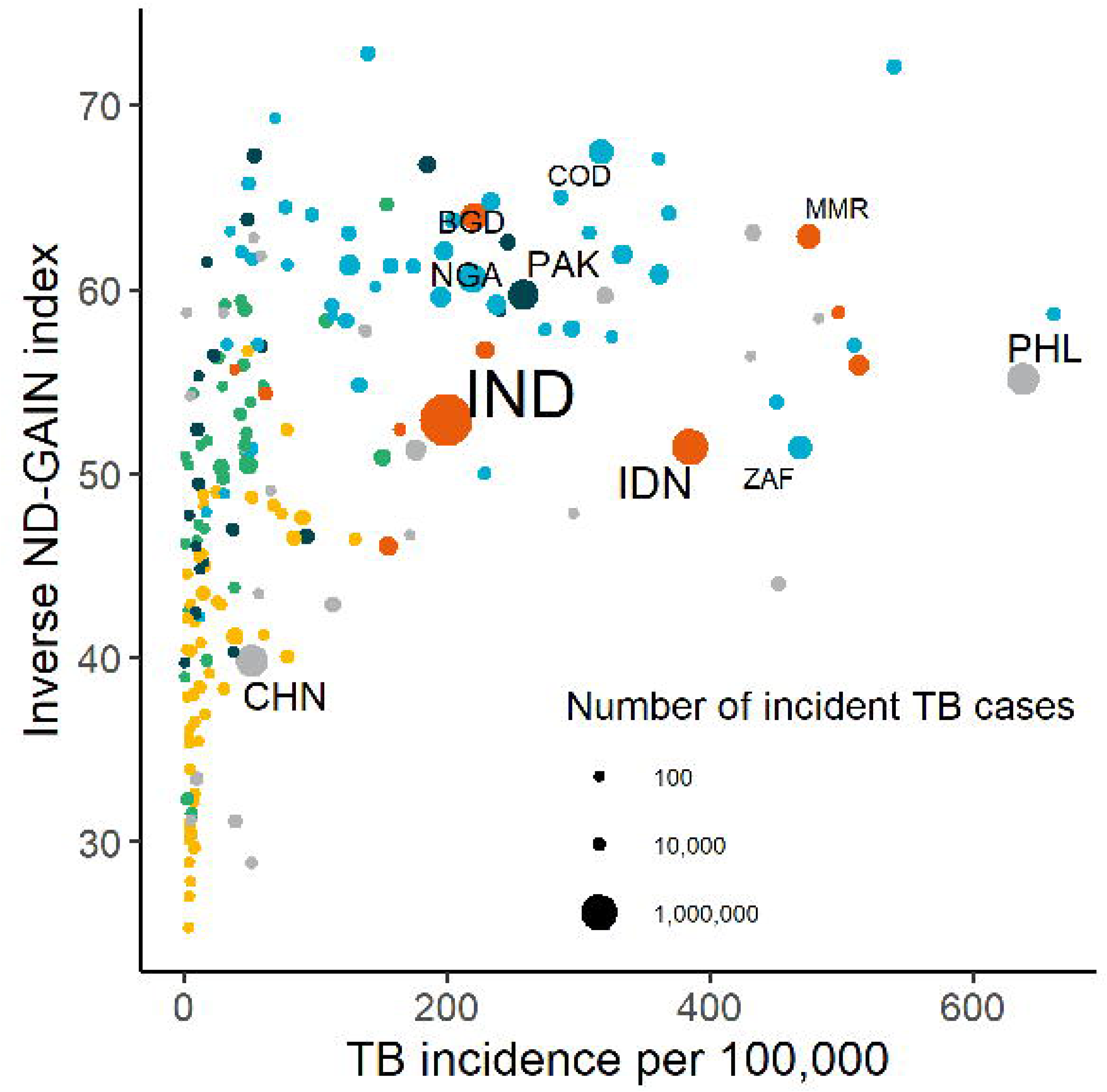
Comparison of TB burden and vulnerability to climate change across 215 countries and territories for different indices. A higher index value represents a poorer performance, where indices include (a) INFORM, measuring the risk of humanitarian crises that could require international assistance, (b) World Risk Index, assessing the risk of humanitarian disaster caused by extreme natural events and the negative effects of climate change, and (c) ND-GAIN, combining both readiness and vulnerability to climate change. Colors indicate World Health Organization regions; blue= African Region, light green= Region of the Americas, dark green= Eastern Mediterranean Region, yellow= European Region, orange= South-East Asian Region, gray= Western Pacific Region. ISO3 codes indicate the top 10 TB burden countries by number of incident TB cases.

In response to this gap, the WHO Global TB Programme commissioned the creation of an analytical framework outlining potential causal relationships between climate change and the TB epidemic, and research gaps to facilitate evidence-building for action. Here, we describe the creation of this framework and its comparison to existing evidence, identify research domains in the area of climate change and TB critically lacking in evidence, and suggest example entry points for intervention.

## Methodology

Development of this framework followed an iterative review process. We first convened an internal working group of TB and climate researchers to develop key questions informing the creation of a preliminary analytical framework, drawing on existing literature, systematic reviews and WHO publications on TB determinants, and wider discussions with key informants and experts. WHO then convened a multi-stakeholder consultation, bringing together representatives of countries, civil society, researchers and public health practitioners, on the Impact of Climate Change on the TB response, where attendees reviewed the analytical framework, supporting evidence, and research gaps. The framework was updated and refined following this consultation meeting.

Our internal working group first identified the principal social and health determinants of TB and conceptualised how these affect different aspects of the TB epidemic and response. We then selected which of these were likely to be sensitive to the effects of climate change, based on previous reviews(10,11), and hypothesised the causal mechanisms through which this might occur. This included identifying relevant climate factors (e.g. changing rainfall patterns or land degradation), the pathway of influence (e.g. socioeconomic changes or migration), and how this might affect TB (e.g. increasing transmission or worsening health outcomes).

After developing and visualizing these hypothesized causal relationships in a preliminary analytical framework, we undertook a narrative literature review. This gathered and synthesized the best available evidence to support, reject, or refine our hypotheses, focusing on three examples of critical pathways: i) migration and displacement, ii) food and water insecurity; and iii) health system disruptions. These example pathways, which are described further below and are termed climate/health links from hereon, were selected based upon:

1. the plausibility of their relationship to climate change, i.e., to what extent a particular pathway was hypothesized to be sensitive to the effects of climate change;
2. their plausibility and importance for TB, informed by their relevance to key WHO End TB Strategy(16) indicators and social and health determinants of TB emphasized by WHO; and
3. the likely availability of existing data and other types of evidence, as well as their amenability for future analysis.

Although each climate/health link could potentially influence multiple consequences for TB, including TB infection, TB disease and TB outcomes, we tested the framework by focusing our literature review on a single TB-related consequence for each climate/health link. For example, we reviewed the effect of health system disruptions on TB outcomes, but did not conduct a comprehensive review of other potential consequences associated with the same climate/health link, such as an increase in transmission due to delayed diagnoses, or higher TB disease rates resulting from reduced preventive treatment coverage.

### Search strategy and selection criteria

We undertook six searches of the MEDLINE database: three investigating the effect of climate change on each climate/health link; and three investigating the effect of each climate/health link on the consequence for TB hypothesized as the primary pathway for that link. Our search strategy (Annex 1) combined searching abstracts and titles for key words with Medical Subject Heading (MeSH) terms. For example, to generate evidence linking climate change to changing TB infection because of migration and displacement, one search combined the concepts: *1) Climate change* with *2) Migration and displacement,* and *9) Study type*; and a second combined *2) Migration and displacement* with *5) TB, 6) Infection*, and *9) Study type.* We further increased our body of evidence by checking references of included articles, citation forward searching (checking for articles that cited included articles), and consulting key experts to obtain information on recent or ongoing studies not captured in our searches. Given the broad scope, our search strategy was refined iteratively, including by identifying MeSH terms and key words for relevant articles and adding them to our search terms if not already included.

We screened titles and abstracts returned by the searches and accessed full-text versions for those potentially eligible for inclusion, including studies in human populations published since 2000 in English. For pragmatic reasons, only evidence from modelling studies or pre-existing reviews (including systematic reviews, meta-analyses, scoping reviews, and narrative reviews) was included. Individual studies, case reports, and perspectives were considered only when no reviews or modelling studies could be found, or where they provided a novel perspective. Key data from included articles were extracted into a standardised form. We then conducted a narrative synthesis to summarise key findings relevant to each topic and used this to adjust and refine the analytical framework towards its final state, including highlighting key areas critically lacking in evidence. This was an iterative process, further informed by discussion with other experts and key stakeholders.

### Analytical framework

The resulting framework is presented in Figure 2. At the highest level, elements are captured describing changing **climate factors**. These cover key examples, which are not exhaustive, such as changing temperatures and rainfall (leading to, for example, increased duration and frequency of droughts or extreme heat), rising sea levels and warming of oceans, and extreme weather events (such as flooding, storms, fires, droughts and extreme heat). Factors are incorporated at a range of timescales, including whether the effects are expected to be visible in the near or longer-term future, as well as how long a given effect might last. For example, extreme weather events have already been widely recorded, and generally result in an immediate but (per event) shorter-term effect. In contrast, sea level rises may currently be a more distal prospect, but one which will likely have a longer-term effect.

**Figure 2.**
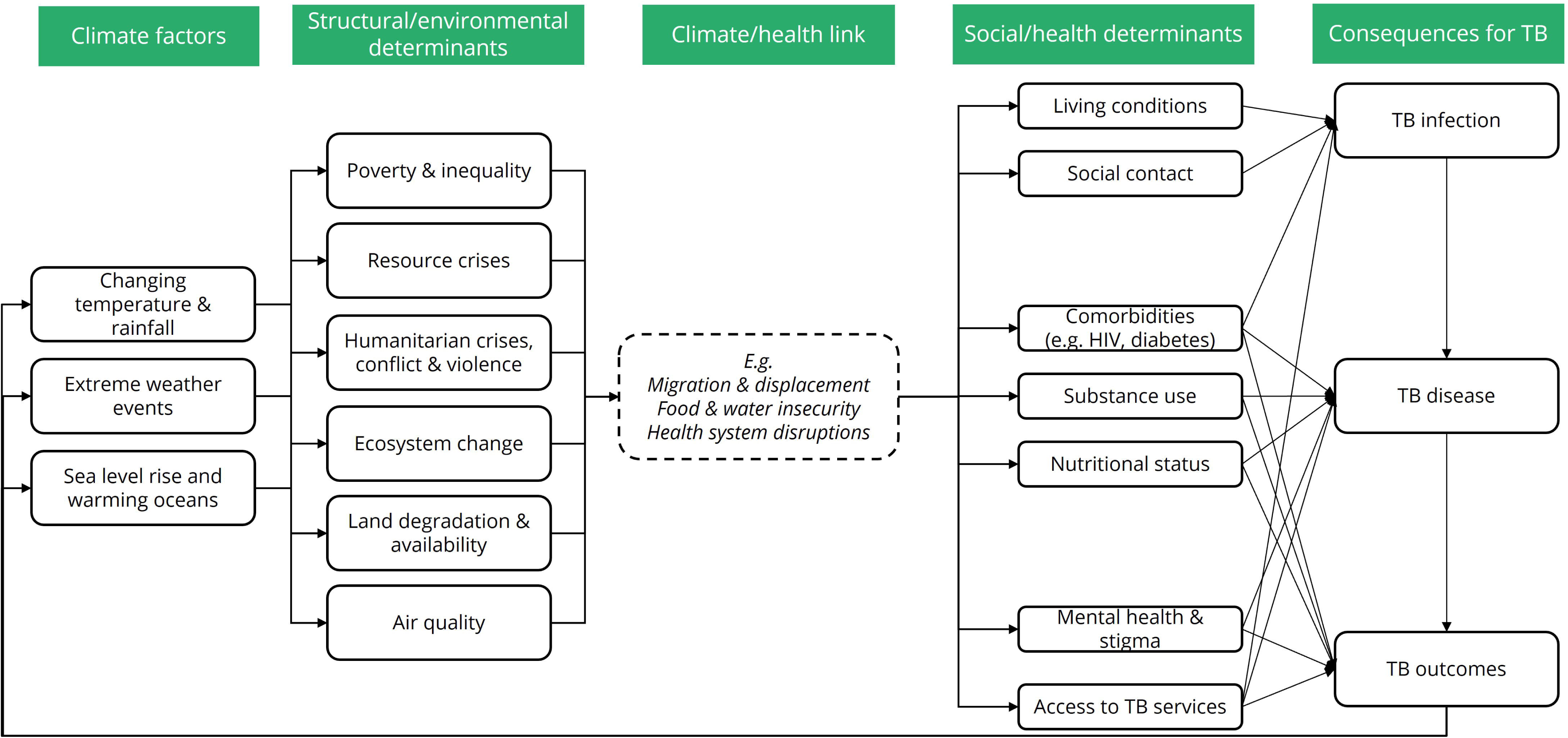
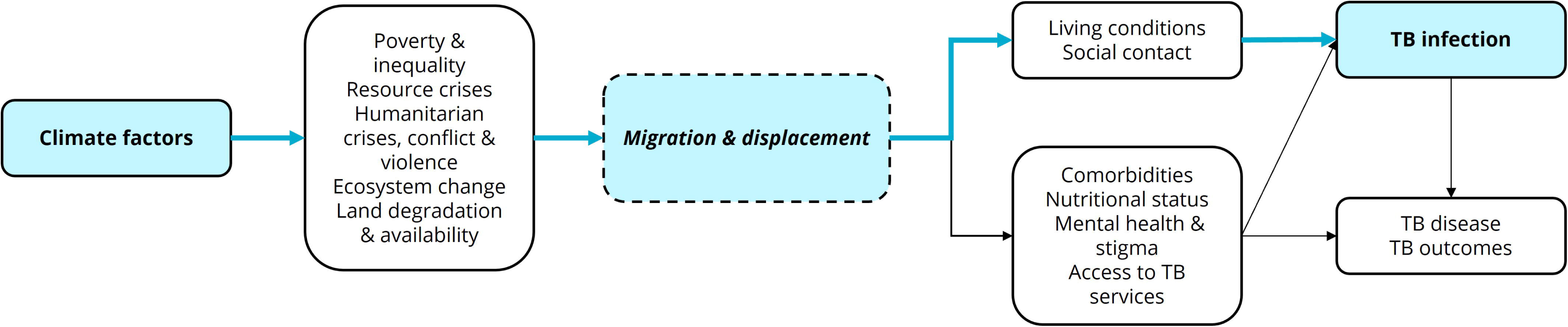
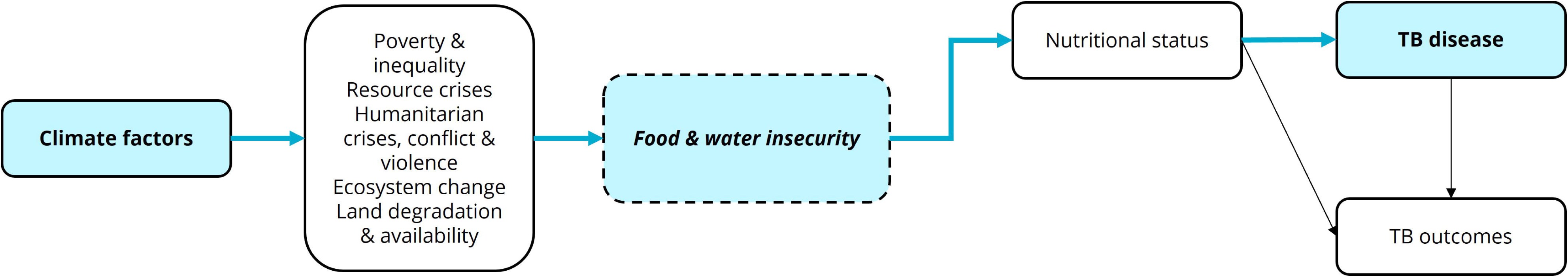
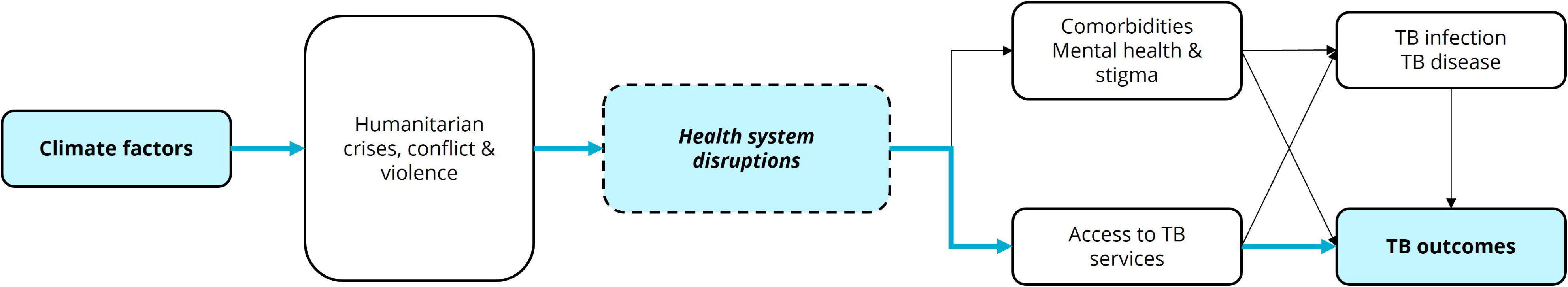
(a) Analytical framework linking climate change to consequences for TB via a range of climate/health links. Example climate/health links considered here include (b) migration and displacement, (c) food and water insecurity, and (d) health system disruptions. Blue arrows indicate pathways included in literature reviews.

As a direct result of these climate factors, a variety of **structural and environmental determinants** of TB are potentially affected. These cover examples such as levels of poverty and inequality (both within and between countries); resource crises (such as fuel, housing, materials and other resources); humanitarian crises, conflict and violence over resource competition; ecosystem change (including changes in seasonality, and in the living and nonliving components of ecosystems); land degradation and availability (including changes to vegetation and available farmland); and air quality.

Changing structural and environmental determinants are then linked to health via a series of **climate/health links**. Three prioritised examples of links are outlined in the sections below and in Table 1; migration and displacement, food and water insecurity, and health system disruption. Each link completes a direct causal pathway through which changes in structural and environmental determinants drive changes in exposure to social and health determinants.

**Table 1.** Examples of prioritised climate/health links.

| **Climate/health link** | **Postulated principle climate factors** | **Postulated structural/ environmental determinant pathway of influence** | **Postulated social/ health determinant pathway of influence** | **Primary postulated consequence for TB** |
| --- | --- | --- | --- | --- |
| Migration and displacement | Changing temperature and rainfall, extreme weather events, sea level rise and warming oceans | Poverty and inequality, resource crises, humanitarian crises, conflict and violence, ecosystem change, land degradation and availability | Changing social contact and living conditions | Increased risk of TB infection, via increased exposure and susceptibility |
| Food and water insecurity | Changing temperature and rainfall, extreme weather events, sea level rise and warming oceans | Poverty and inequality, resource crises, humanitarian crises, conflict and violence, ecosystem change, land degradation and availability | Changing nutritional status | Increased risk of TB disease, via increased likelihood of TB progression |
| Health system disruptions | Extreme weather events | Humanitarian crises, conflict and violence | Changing access to TB services | Poorer outcomes, via increased vulnerability to TB and reduced diagnosis and care |

**Social and health determinants** cover examples such as living conditions and housing (affecting factors such as overcrowding and ventilation); changing social contact as individuals move within and between countries; and changes in prevalence and management of comorbidities including (but not limited to) HIV and diabetes, changes in nutritional status, mental ill health and stigma, and provision of and access to TB prevention and care services.

Lastly, social and health determinants are explicitly linked to **TB consequences**, where they affect;

1. the likelihood of exposure to and susceptibility to TB infection;
2. the risk of progression to TB disease; and
3. the extent of vulnerability to TB outcomes (including short- and long-term morbidity, disabilities, mortality, acquisition of drug-resistance, and psychological and financial consequences).

These elements can impact progress towards the goals and targets of the WHO End TB strategy. Increased TB exposure and susceptibility may drive higher transmission, leading to greater infection rates, disease burden, and associated health and social costs. Similarly, TB disease progression influences health outcomes, onward transmission and incidence, and TB-related socio-economic consequences. Moreover, worsening inequalities can exacerbate health disparities, including mortality and TB-related catastrophic costs.

TB care and prevention programmes may also contribute to climate change, creating a feedback loop within the framework. The healthcare sector more generally is estimated to account for approximately 5% of global emissions(17), and, when specifically considering TB, there are multiple sources of emissions across the TB care cascade(18,19). These include the energy and material inputs required for TB diagnostics and treatment, transportation emissions from patient visits and sample transport, and the medical and biological waste generated as part of providing care.

Due to the interconnectedness of potential pathways, many of which are cyclic or mutually reinforcing, the framework is not intended to be exhaustive, but seeks to capture the pathways with significant implications for the TB epidemic and response, and those that are amenable to analysis and actionable from a policy perspective. As part of this work, the pathways for the three climate/health links described above were reviewed in detail (Table 1). Two other potential pathways, not reviewed in detail for pragmatic reasons, but which merit further attention, are air pollution and meteorological factors (Panels 1 and 2).

Importantly, these pathways often create compounding vulnerabilities. For example, displaced populations frequently have reduced access to adequate nutrition and healthcare; those living in poverty who are experiencing food insecurity often concurrently have limited access to healthcare and higher displacement risks; and when extreme weather events cause disruptions to health systems, they typically also cause displacement and disruptions to food security.

### Migration and displacement

#### Evidence on climate change and migration and displacement

Climate change and migration and displacement has an extensive history of research(3), with multiple existent frameworks(20–23) and a wealth of examples of how climate change might drive population movement both between and, more commonly, within countries(24–34). In 2023, approximately 20.3 million people were internally displaced as a result of weather-related hazards, while by 2050 that number could increase to 216 million due to slow-onset climate change impacts(35). Several reviews cover circumstances encompassing natural hazards and socioeconomic changes(36–43), including reviews comparing different climate migration models(44–48). Models projecting future migration due to climate change are also numerous. Multiple examples focus on sea level rise and flooding(49–56); although some consider meteorological(57–63), macroeconomic(64,65), and agricultural changes(66,67) and more general frameworks exist(68–72). Given this the IPCC states with high confidence that extreme weather events and variability act as direct drivers of involuntary migration and displacement, and as indirect drivers through deteriorating economic conditions and livelihoods(3). However, patterns of migration due to climate change are likely to be highly context specific and are difficult to project because of the multicausal nature of migration, migration policies, and the scale and nature of any future adaptations. Nevertheless, it is striking that the IPCC states that under all global warming levels there are areas of the world which will become unsafe/uninhabitable, many of which are currently densely populated and have a high TB burden(3).

#### Evidence on migration and displacement and TB infection

There is also a large body of evidence linking migration (particularly forced migration and displacement) to TB risk(73,74). While no routine estimates of burden are published, reviews suggest a multi-fold increased risk of TB for displaced persons(75,76). The literature focuses principally on the additional TB disease burden and poor treatment outcomes (partly because of reduced access to care) experienced by displaced people or those in crisis situations(75–79), or on TB disease screening in migrants from high to low TB burden settings(73,80–89). This makes it difficult to establish whether any increase in TB disease in migrants compared to non-migrants from the same region is driven by an increase in transmission and infection resulting from changing living conditions and social contact during transit, in camps and asylum settings, or due to progression to TB disease due to poor nutrition or poor access to preventive care because of barriers such as language, legal status, low resources, stigma, or a combination of all of these. While we focus here on TB infection, it is important to highlight that a combination of several factors described above, including underlying health vulnerabilities, play a role in increasing disease risk throughout the migration and displacement journey(79). In the context of human mobility and TB infection, a review of the impact of conflict on infectious disease found an increase in TB transmission due to displacement(90). A separate review of migrants to low TB burden settings identified case studies where transmission during transit led to geographically widespread clusters(73). Another review comparing transmission in foreign- and native-born communities(91), found that TB in foreign-born populations did not have a significant impact on TB among native populations in Europe. One individual study compared TB infection prevalence, finding no evidence for a link to living in a disaster area but some evidence for a link to overcrowded living conditions(92), while a modelling study considering rural-urban migration in China identified the important role of migration in transmission(93). While multiple reviews identify a high risk of TB infection among migrants across a range of settings, they frequently do not provide direct comparisons with populations of origin to assess changes in transmission risk(78,94–97).

### Food and water insecurity

#### Evidence on climate change and food and water insecurity

Climate change and food and water insecurity likewise have long been known to be intrinsically linked(3), and were the focus of a recent special IPCC report(98). Approximately 733 million people faced hunger in 2023, equivalent to one in eleven people globally and one in five in Africa, and if current trends continue 582 million will still be chronically undernourished by 2030(99). Short-term disruptions to food systems due to climate-driven extreme weather events or other disasters such as earthquakes and conflict have been shown to have a direct effect on food and water security as well as nutritional status(99–113), with exposure to such acute events potentially leading to lasting consequences(114,115). Meanwhile, longer-term effects of changing temperatures and precipitation on crop yields, grassland quality, and oceans (through warming and acidification) have already been observed to negatively impact agricultural and aquaculture productivity, with significant future impacts on food security expected(3,116–123). Furthermore, a large body of literature exists reviewing the effects of climate change directly on nutritional status and associated health outcomes(124–131), as well as projecting longer-term effects due to changing calorific availability and diets (132–153), including because of increased food costs. The focus of many of these studies is on malnutrition in children, including stunting and wasting, or on obesity and overweight in adults. Several studies were also found on the effect of climate-induced food insecurity on birth weight, which were not included here. Importantly, although the effects of climate change on food and water insecurity are likely to affect everyone to some extent, they are likely to disproportionately affect high TB burden countries with underlying vulnerabilities in their food systems(3).

#### Evidence on food and water insecurity and TB disease

Abundant evidence exists linking food and water insecurity to TB via the pathway of undernutrition. Undernutrition doubles the risk of TB disease, and nearly 10% of TB cases globally are attributable to undernutrition(6), although this proportion is likely higher in many high TB burden countries(154). Most studies use body mass index (BMI), a widely- and easily-used indicator of nutritional status which has been demonstrated to have a log-linear relationship with TB incidence in a range of settings and populations(155,156). Several other systematic reviews and meta-analyses further confirm the association between increased TB disease risk and undernutrition, including in people living with HIV or people with diabetes (157–160). The reverse has also been shown to be true; interventions addressing food insecurity such as provision of food baskets have been shown to reduce TB disease risk(161), In addition, modelling studies have project large reductions in TB burden if undernutrition is addressed(162–164), and the reverse if it worsens(165). Other important pathways exist by which food and water insecurity affect TB consequences. Chief amongst these is an increase in poor TB treatment outcomes associated with undernutrition(166–169), whereas improvements in nutritional status likely improve outcomes(170).

### Health system disruptions

#### Evidence on climate change and health system disruptions

Around 3.5 billion people live in areas highly vulnerable to climate change, with direct consequences for their access to healthcare services(171). There is a substantial body of evidence linking climate change and health systems disruptions(3,79,171–173), demonstrating how climate events compromise healthcare infrastructure, disrupt service provision, and strain the health workforce. Existing literature on health system disruptions predominantly examines the impact of extreme weather events (particularly flooding and storms) on healthcare service delivery, with several reviews emphasizing disruptions to chronic disease management(174–180). One review focuses specifically on challenges faced by the healthcare workforce and how to mitigate these(181), while another evaluated preparedness of hospitals for disasters(182). Further studies from oncology and maternal health highlight how extreme weather events affect access to health services, particularly for vulnerable populations(183–186). These studies provide lessons on changes in healthcare utilization between affected and unaffected communities, and by socioeconomic position(187–190). Studies concerning other natural disasters such as earthquakes and volcanic eruptions were not included in our review but may still provide relevant insight. Beyond natural disasters, emerging evidence highlights additional climate-related disruptions to healthcare. For example, a modelling study demonstrated how emergency department visits may change due to increasing temperatures(191), while another review evaluated the effects of economic recessions (not necessarily climate-induced) on healthcare(192).

#### Evidence on health system disruptions and TB outcomes

Much of the evidence on the effects of health system disruption on TB focuses on the recent COVID-19 pandemic, with a strong emphasis on reductions in TB case notifications, a proxy indicator for the number of people reported to have accessed care. These disruptions alone are estimated to have led to nearly 700,000 excess TB deaths between 2020 and 2023(6). Several reviews have collated evidence on the effects of disruptions associated with the pandemic on the TB care cascade(193–197), with some explicitly considering TB treatment and outcomes(6,194,197–200). Meanwhile, modelling studies projected the possible consequences for multiple settings, finding significant increases in incidence and mortality(6,201–206). Due to the nature of the disruptions, most studies combined the effects of disruptions to service delivery and human resources together with disrupted supply chains, with little evidence characterising the effect of disruptions to infrastructure and technologies, energy or sanitation. There is also little evidence on the effect of disruptions on other outcomes, such as disease severity or catastrophic costs. Outside of the pandemic, two recent reviews of the effect of conflict on TB also identified studies from a range of settings, again focused primarily on diagnostic delay and treatment interruption(79,90). Meanwhile, an earlier modelling study evaluated the effect of an Ebola outbreak, finding a reduction in TB diagnosis and treatment success (207).

## Discussion

Through the development of this analytical framework and by undertaking comprehensive literature reviews, we have demonstrated that the effects of climate change on the TB epidemic are likely to be mediated through multiple pathways, and have the potential to be highly consequential. Specific effects will vary by the magnitude of the climate hazard(s), the vulnerability of communities to their effects (e.g. due to differences in underlying TB determinants), and the capacity of communities to adapt (determined by factors including income, living conditions, and access to healthcare and social protection). Importantly, people affected by TB are particularly vulnerable to the effects of climate change because they are already disproportionately poor, undernourished, and may have comorbidities such as HIV. This hinders their ability to adapt effectively, perpetuating inequality and injustice.

Outlined in Table 2 are a number of research gaps evident from the literature review and a consultation meeting on the Impact of Climate Change on the TB Response convened by WHO, which focus specifically on the three climate/health links described previously. Overall, whilst evidence exists demonstrating the relationship between climate change and each climate/health link (e.g. climate change and food and water insecurity); and between each climate/health link and TB (e.g. food and water insecurity and TB disease), there were no studies directly quantitatively linking climate change and TB, via any of the described climate/health links or otherwise.

**Table 2.** Example research domains and questions to more effectively and comprehensively characterise the effects of climate change on TB, and how to mitigate these effects and adapt and build resilience to them.

| Research domain | Specific example questions |
| --- | --- |
| *Climate change and TB*<br>No evidence exists directly linking climate change to TB. There is an urgent need to generate such evidence, as well as to identify prioritised adaptation and mitigation measures for climate-resilient TB services with a minimal environmental footprint. | Are the effects of climate change on TB likely to be similar in high and low burden settings, and to what extent could climate change impact the global epidemiology and distribution of TB (e.g. could the effects change a country from a low burden to a high burden setting?) |
|  | How do the effects of climate change on TB vary among different population groups (e.g. by gender, or people in extreme poverty)? |
|  | Which other climate/health links (e.g. air quality and extreme heat) might have significant causal effects on TB, beyond those currently explored? |
|  | How does the environmental footprint of TB prevention and care (e.g. diagnostics and the treatment process) contribute to climate change, and what sustainable practices can be introduced to minimize this impact? |
|  | What adaptation and mitigation measures (e.g., improved housing, social protection, resilient healthcare systems) should be prioritized to build climate-resilient TB services, particularly in resource-constrained settings? |
| *Climate change, migration and displacement, and TB infection*<br>Evidence linking climate change to migration and displacement has focused on population movement in | Do existing theoretical and operational frameworks adequately capture the complex, multifaceted effects of climate change on migration, including acute displacement and longer-term population shifts? |
|  | How will overcrowding, poor living conditions and increasing urbanization driven by climate change affect TB transmission |
| the longer term, particularly due to sea level rise, with less evidence on when this is likely to lead to overcrowding, increases in urbanization, or poorer living conditions.<br>Evidence linking migration and displacement to TB infection has focused on TB disease among migrants in low-burden settings, with limited data on how infection risk changes and the effect on refugees, asylum seekers, and internally displaced people. | risks and health outcomes (including mental health) in displaced populations? |
|  | What are the barriers to healthcare access for displaced populations affected by climate change, and how do these barriers influence TB detection, treatment adherence, and outcomes? |
|  | What social and economic determinants related to climate change and displacement (e.g., unemployment, poverty, food insecurity) most strongly influence TB risk in displaced populations? |
|  | How does migration type and experience (e.g. internal displacement or asylum seeker etc.) alter transmission dynamics, risk of exposure, and health outcomes? |
|  | Can studies quantify the effect of different forms of climate-induced migration and displacement on TB transmission risk, particularly in high-burden and climate-vulnerable areas? |
| *Climate change, food and water insecurity, and TB disease*<br>Evidence linking climate change to food and water insecurity has focused on children and the overweight, particularly due to changing food availability and diets, with more limited data on undernutrition in adults and the effects of extreme weather events and acute nutritional shocks.<br>Evidence linking food and water insecurity to TB disease has focused on BMI as an indicator, with more limited data from high TB burden settings, in populations with a | How will the growing frequency and intensity of extreme weather events and their effects on food and water insecurity subsequently affect susceptibility to TB infection, disease progression and TB outcomes, and what are the most effective forms of nutritional support and anthropometric markers to deploy in these settings, both in the short and long term? |
|  | How does the relationship between BMI and TB incidence vary across different populations, including children and older people and those with comorbidities such as HIV infection, alcohol use disorders, or end-stage renal disease; and does the type of weight loss (acute vs. chronic) or BMI distribution in a setting influence TB risk, even in the absence of malnutrition? |
|  | Can additional nutritional indicators—such as hemoglobin levels, body composition (via bioelectrical impedance |
| particularly high prevalence of severe underweight, for children and adolescents, or for other nutritional indicators. | analysis), point-of-care micronutrient testing, and measures of dietary diversity and quality—offer more precise evidence of vulnerability to TB disease, beyond BMI alone, and what are the most appropriate nutritional metrics for different contexts? |
|  | Can models quantify the added impact of increased food and water insecurity driven by climate change on people vulnerable to TB? |
| *Climate change, health system disruptions, and TB outcomes*<br>Evidence linking climate change to health system disruptions has focused on extreme weather events and service delivery/access, with little evidence on more gradual disturbances from repeated shocks or climate-driven economic slowdowns, or the effect on supply chain disruptions, infrastructure, energy, and sanitation.<br>Evidence linking health system disruptions to TB outcomes has focused on the COVID-19 pandemic and case notification, with limited evidence from other forms of disruptions, or for a wider range of health and socio-economic outcomes. | How will factors beyond extreme weather events, such as prolonged economic crises or extreme heat, contribute to health system disruptions, and how will these affect TB service delivery? |
|  | In what ways will climate change impact essential health service functions (e.g., supply chains, infrastructure, energy availability, health workforce, sanitation) critical to effective TB response? |
|  | What are the long-term consequences of health system disruptions caused by climate change on TB-related health outcomes, including the risks of chronic lung damage, the acquisition of drug-resistant TB strains, and financial burden on affected populations? |
|  | Can experience from previous disruptions be used to quantify the effects of acute climate events (e.g., floods, droughts) on TB outcomes as a result of disrupted service delivery, including disruptions to TB diagnostics, supply chains, and treatment delivery? |

The available evidence demonstrates that efforts to mitigate the effects of climate change on the TB epidemic must adopt a multisectoral approach that addresses underlying TB determinants, and is also responsive to the unique needs of vulnerable populations, especially migrants and displaced populations. Immediate actions should include meeting core obligations under the right to health by ensuring universal health coverage and establishing a social protection floor for all individuals. This requires mobilizing adequate investment to build resilient health systems and to mitigate adverse economic and non-economic fallouts for vulnerable populations. Using the framework to identify relevant entry points for the three climate/health links discussed here, Table 3 outlines some examples of specific measures, which can be mapped to where in the pathway they intervene. Further research is urgently required to develop a prioritised set of feasible, practical and impactful interventions ultimately aiming to address both the short- and long-term effects of climate change.

**Table 3.** Examples of cross-sectoral intervention entry points.

| Climate/health link | Examples of interventions | Potential sectors involved in intervention |
| --- | --- | --- |
| Migration and displacement | Migration-sensitive TB screening programmes; infection control and prevention measures; housing programmes; equitable and affordable access to | Health, migration, housing/social protection |
|  | health care services |  |
| Food and water insecurity | Sustainable agriculture and water resource management interventions; strengthened food supply chains to ensure accessibility and affordability; social protection programs targeting vulnerable populations; cash and in-kind transfers; targeted nutritional interventions to address malnutrition and micronutrient deficiencies; | Health, agriculture, finance/social protection |
| Health system disruptions | Health system and supply chain strengthening; mobile care and digital health systems ensuring continuity of TB services and treatment support; decentralisation of care and training health professionals to manage climate-sensitive health challenges | Health, environment |
| Planning and coordination in countries most affected by TB | Integration of TB prevention and care into national plans of action for climate mitigation and adaptation; participation of people affected by TB, particularly those most vulnerable to climate impacts, in policy-making processes related to climate mitigation and adaptation | Health |

To achieve these objectives, States must first recognize the interlinkages between TB and climate change, and ensure the End TB Strategy is implemented synergistically with other global agreements, including the Paris Agreement, the Sendai Framework for Disaster Risk Reduction 2015–2030, the 2030 Agenda for Sustainable Development, and the Global Compacts for Migration and Refugees(208–212). National TB programmes can serve as entry points by monitoring the effects of climate change to build evidence and support decision-making, and foster cooperation with national adaptation policies and institutions. Civil society organizations, particularly those representing TB-affected communities, play an important role in amplifying the voices of those affected by TB within climate mitigation and adaptation initiatives, advocating for equitable policies, and mobilizing resources. These local efforts must be supported by international institutions and collaborative mechanisms to exchange best practices, share data, strengthen evidence, and propose effective mitigation strategies, drawing insights from other health contexts where applicable. Lastly, the effect of the End TB Strategy on climate should also consider how to mitigate the environmental impacts for all the inputs required to ensure and deliver person-centered prevention, treatment and care, ultimately aiming to encourage TB programmes to adopt less carbon-intensive measures and move forward carbon-neutral TB programmes.

In conclusion, we have shown how evidence supports the existence of causal links between climate change and TB, and how further evidence is urgently needed to quantify the extent of impact on TB. Furthermore, TB needs to be included in climate risk adaptation and mitigation programmes, and climate-resilient funding and response mechanisms. As our work shows, climate change is already hindering progress in the fight against TB, and only through comprehensive action can we prevent it becoming a barrier to ending the global TB epidemic.

## Data Availability

All data produced in the present study are available upon reasonable request to the authors

## Contributors

DB, RGW, RMGJH, MZ, NG and CFM conceived the study. MJS, DB and CFM conceived and designed the framework. All authors critiqued the framework and study approach. MJS, DB, PYK, LG and CFM conducted the literature review. MJS and CFM wrote the first draft of the article. All authors revised and edited the submitted Article. MJS and CFM had full access to all the data in the study and had final responsibility for the decision to submit for publication.

## Acknowledgements

This work was funded by the WHO (APW 203462345). MJS is also funded by NIHR through a Clinical Lectureship. CFM is also funded by BMGF (TB MAC OPP1135288, INV-059518) and NIH (R-202309-71190). RAC is also funded by BMGF (INV-001754), and NIH (G-202303-69963, R-202309-71190). RGW is also funded by the Wellcome Trust (310728/Z/24/Z, 218261/Z/19/Z), NIH (1R01AI147321-01, G-202303-69963, R-202309-71190), EDTCP (RIA208D-2505B), UK MRC (CCF17-7779 via SET Bloomsbury), ESRC (ES/P008011/1), BMGF (INV-004737, INV-035506), and the WHO (2020/985800-0). RMGJH is also funded by the Wellcome Trust (310728/Z/24/Z) and NIH (R-202309-71190). JMP is also funded by the Wellcome Trust (305644/Z/23/Z).

## Panel 1. Air pollution

Air pollution is an important climate/health link that is not reviewed in detail in this document but may have particular relevance for respiratory diseases such as TB.

Nearly the entire global population breathes air that contains pollutants exceeding the levels recommended by WHO guidelines(213). These pollutants, including particulate matter (PM2.5 and PM10) and sulphur dioxide, are a known cause of major cardiovascular and respiratory diseases, including cancer. Because they also impair lung defence mechanisms and may modulate the immune response, chronic exposure to these pollutants may increase susceptibility to TB infection, progression to TB disease, and risk of adverse TB outcomes. Indeed, the association between exposure to *household* (indoor) air pollution (from burning solid fuels for heating or cooking) and TB incidence has been established for some time(214). For *ambient* (outdoor) air pollution, a recent systematic review demonstrated an association with increased TB incidence(215), but not with hospital admission or mortality, and highlighted the low availability and quality of evidence. Further research is therefore required in this area, including research to establish the exact causal pathways (i.e. whether air pollution increases susceptibility to TB infection, progression to TB disease, or both).

Importantly, ambient air pollution and climate change are strongly interconnected. Many of the causes are the same, principally the burning of fossil fuels and deforestation/agricultural practices, and there are also multiple feedback loops(216). Drier and hotter conditions can increase ambient air pollution directly through increased photochemical production and air stagnation events, and indirectly through wildfires(217). Many air pollutants (especially black carbon) directly contribute to global warming by absorbing solar radiation and trapping heat in the atmosphere(218). Climate change may also increase exposure to household air pollution if extreme heat or weather drives people to spend long periods indoors. These interconnections mean that interventions to address either air pollution or climate change will generally, although not universally, have beneficial effects on the other.

## Panel 2. Meteorological factors

The associations between specific, measurable meteorological factors such as temperature, humidity, and precipitation and TB have been the subject of some research, primarily in Asia, and these factors have been proposed as potential indicators that could be used to predict future changes in TB incidence(219–221). Results, however, may be inconsistent. For example, some studies suggest that increases in average temperature are associated with increased TB incidence(222–224), whilst others report that increased temperatures are protective(225–228). A recent systematic review and meta-analysis of meteorological factors and TB found that TB risk was positively correlated with precipitation exposures but not average temperature, humidity, air pressure, or sunshine duration(229). Another meta-analysis on ecological-level factors and TB found higher humidity and precipitation were associated with increased TB incidence, whilst higher wind speed was associated with reduced TB incidence(230). It has also been posited that antimicrobial resistance may independently increase as temperature increases(231).

Importantly, these associations alone provide little insight into the pathways through which such meteorological factors might impact TB. Relatedly, seasonal variations in TB case notifications have been demonstrated in multiple settings(232–235), with explanations proposed ranging from seasonal-related migration (mostly for economic opportunities); to indoor crowding (because of cold weather or extreme heat); and vitamin D deficiency in winter manifesting as a spring peak in notifications. Ultimately, the relationship between meteorological factors and TB is likely to be highly context-specific, varying by geographical region, local climate patterns, and socioeconomic conditions, and further research is required in this area.

## Annex

**Table 1.**
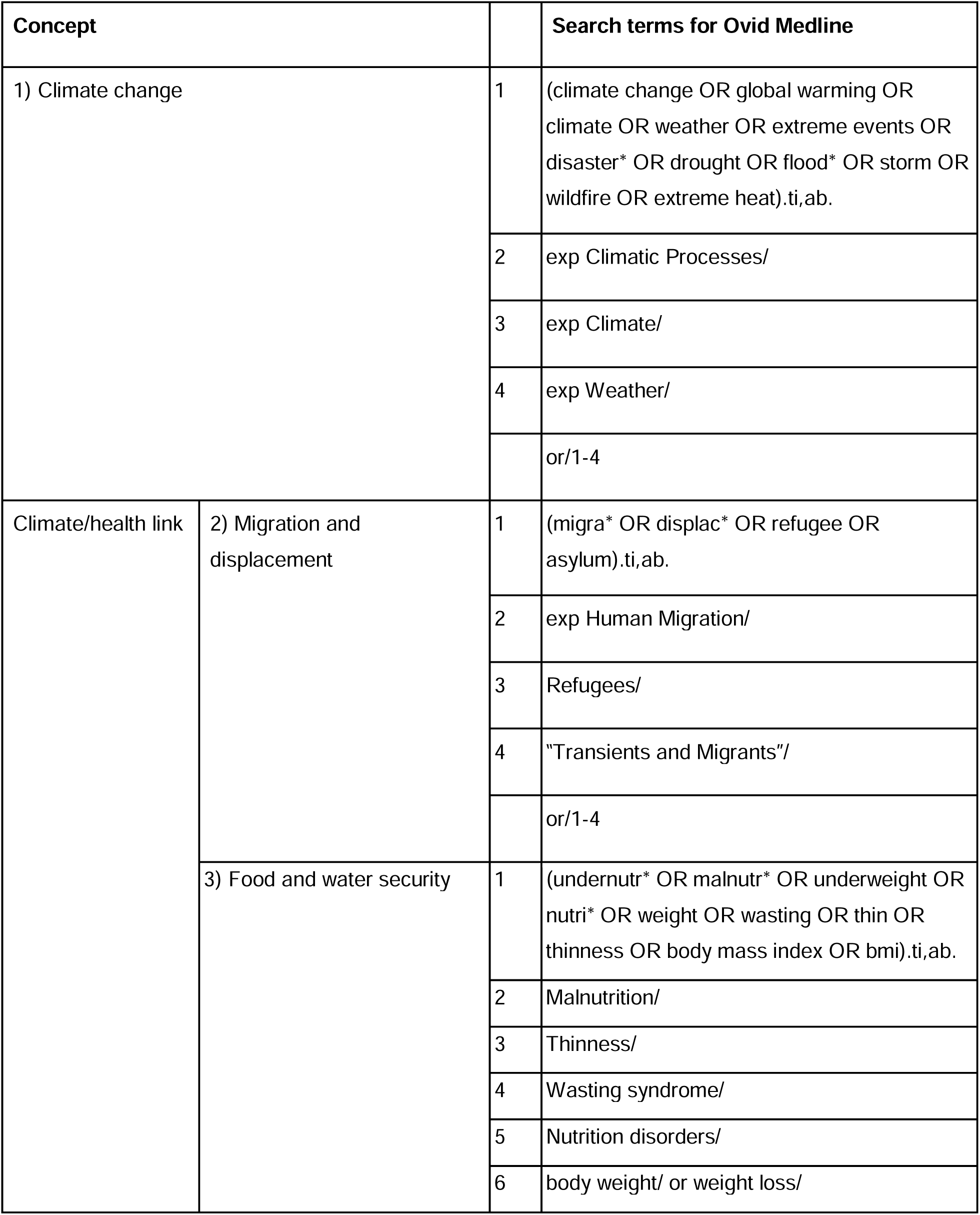
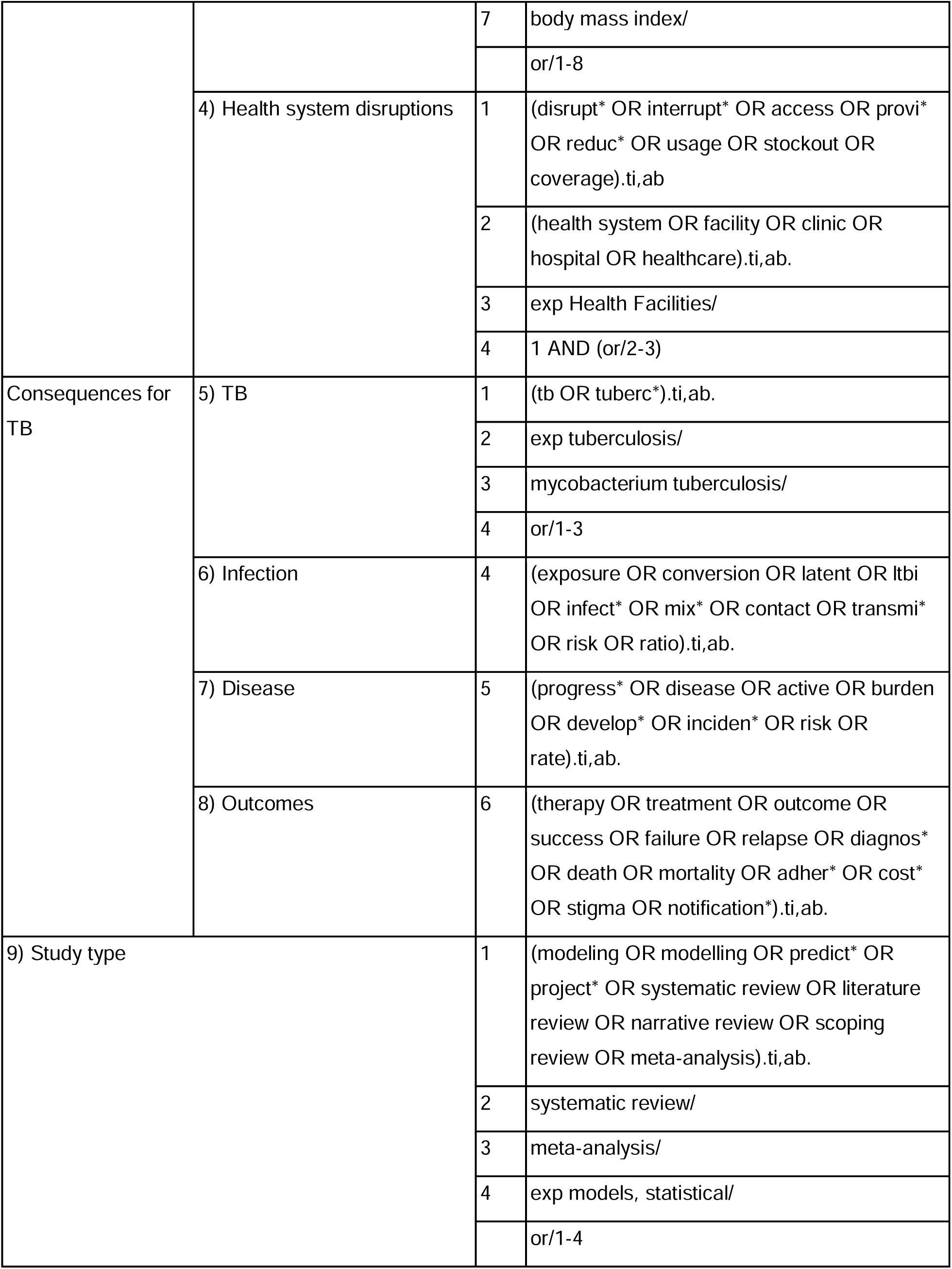
Search strategy terms.

**Table 2.** Number of records returned by searches.

| Search | Number of articles |
| --- | --- |
| Climate change AND migration and displacement | 2,324 |
| Migration and displacement AND TB infection | 356 |
| Climate change AND food/water insecurity | 6,515 |
| Food/water insecurity AND TB disease | 1,412 |
| Climate change AND health system disruptions | 2,574 |
| Health system disruptions AND TB outcomes | 1,459 |

